# Exercise-induced hypoalgesia following proprioceptive neuromuscular facilitation and resistance training among individuals with shoulder myofascial pain: a pilot study

**DOI:** 10.1101/2022.06.28.22276990

**Authors:** Zi-Han Xu, Nan An, Zi-Ru Wang

**Affiliations:** School of Sport Medicine and Rehabilitation, Beijing Sport University, 48 Xinxi Road, Haidian District, Beijing, China

**Author notes:** Corresponding author: Xu, Z.H. Xu, Z.H., An, N. and Wang, Z.R. contributed equally to this manuscript. Registration: This study has been registered in Chinese Clinical Trial Registry. Registration number: ChiCtr202111090819166165. Full trial protocol can be accessed in http://www.chictr.org.cn/showproj.aspx?proj=135425.

**Keywords:** exercise induced hypoalgesia, proprioceptive neuromuscular facilitation, resistance exercise, conditioned pain modulation, myofascial pain syndrome

## Abstract

**Objective:** The present study estimated the effect of proprioceptive neuromuscular facilitation (PNF) and resistance training on exercise-induced hypoalgesia (EIH) and conditioned pain modulation (CPM) among patients with myofascial pain syndrome (MPS).

**Methods:** A total of 76 female MPS patients (aged from 18-30) with a visual analog scale (VAS) score greater than 30/100 mm were enrolled in the study. Participants were randomly assigned into 3 intervention groups, including isometric (n=18), isotonic (n=19) and PNF (n=20) exercises, and 1 control group (n=19) with no intervention. Pressure pain threshold (PPT) and the CPM responses at myofascial trigger point, arm and leg sites were assessed before and after exercise session.

**Results:** There was an increase in PPT and CPM responses at trigger point, arm and leg sites in participants performed PNF and isotonic exercise, while the isometric exercise only increased PPT at leg sites. Compared with control group, both isotonic and PNF group showed greater EIH responses at the trigger points. However, only the PNF exercise significantly improved PPT and CPM responses at arm and leg sites compared to the control group.

**Conclusions:** PNF, isotonic and isometric exercises could lead to local and global EIH effect. The increase in CPM response after PNF and isotonic exercises indicated that the EIH mechanisms of different resistance exercises may be attributed to the enhancement of the endogenous pain modulation through the motor-sensory interaction from the additional eccentric and dynamic muscle contraction.

## Introduction

Exercise therapy has been recommended in many chronic pain treatments and managements^1^, regarding its advantages in efficiency, cost, safety and the exercise-induced hypoalgesia effect (EIH)^2^ in either healthy or individuals suffering pain. Both aerobic and resistance exercises may attenuate pain perception globally by enhancing endogenous analgesia^3^ via descending pain modulation. However, patients with myofascial pain syndromes (MPS) always demonstrate dysfunction in descending pain modulation^4^, which could excite and disrupt pain perception, even lead to attenuation of the hypoalgesia effect induced by maximal and submaximal aerobic^5^ or resistance exercises^6^.

The altered endogenous pain modulation may be the reason for impaired EIH for MPS patients, for which showed both local and global hyperalgesia. Normally, the sufficient noxious or non-noxious input from the C afferent fibers can activate thalamic mediodorsal (MD) nucleus and ventromedial (VM) nucleus^7^ by different thresholds, triggering the descending facilitation (low-thresholds) or inhibition (high-thresholds)^8^ respectively. And the On or Off cells in periaqueductal gray (PAG)^9^, locus coeruleus (LC) and rostral ventromedial medulla (RVM)^10^ projected to the spinal dorsal horn (SDH) can active the NMDA receptors^11^ to facilitate the ascending nociceptive signals, or release the endogenous opioid^12^, 5-HT^13^ and noradrenaline^14^ to inhibit. Meanwhile, the pain-related brain regions including primary motor cortex (M1)^15^, anterior cingulate cortex (ACC)^16^ and insula cortex^17^, et al a can also participate the descending modulation of pain through the motor-sensory interaction. However, the prolonged pain condition including MPS can develop into central sensitization, where the balance between the descending facilitation and inhibition is broken and may deficit the EIH effects.

With respect to the pathway of endogenous pain modulation, it’s believed that the sensory input during the exercise, especially the proprioception and C fibers can play the critical roles in the EIH. Thus, proprioceptive neuromuscular facilitation (PNF) exercise^18^, which can enhance the C fibers^19^ and proprioception^20^ inputs through the combination of various eccentric and dynamic exercises, may have additional positive effects on EIH in people with chronic pain, while the preliminary therapeutic effect of PNF on pain of MPS patients^21^ was also demonstrated. In fact, the type of muscle contraction can affect the EIH in chronic pain isometric exercise could attenuate pain sensitivity in shoulder pain but not in fibromyalgia^22^, while the isotonic exercise performed in no-painful limbs could reduce pain perception in chronic pain of knee^23^. However, few research compared the effects of various resistance exercises on EIH in MPS, and the relationship between exercise type and endogenous analgesia features regarding EIH in MPS is also needed further investigation.

In addition, conditioned pain modulation (CPM)^24^has been applied to evaluate the descending pain modulation in patients with chronic musculoskeletal pain. And the deficit in endogenous analgesia of MPS^25^ was demonstrated by the attenuation of CPM responses. CPM has been shown to predict EIH in patients with knee osteoarthritis receiving bicycling and isometric exercise^26^, while EIH has similar effects compared to CPM in healthy individuals^27^, but the relationship between descending pain modulation and EIH has not been investigated so far in MPS patients intervened by PNF and other resistance exercises.

Therefore, this pilot study aims to compare short-term EIH responses following PNF, isotonic and isometric resistance exercises, and to investigate the relationship between EIH and descending pain modulation determined by CPM in patients with MPS. It was hypothesized that the PNF and all resistance exercise would have a higher EIH response compared to blank control in individuals with MPS. The EIH and CPM response at baseline would be relatively lower in MPS patients with impaired descending pain modulation, but would increase or restore following PNF and resistance exercise intervention performed on the affected areas.

## Methods

This study has been approved by Sports Science Experimental Ethics Committee of Beijing Sport University (ethics approval number: 2021153H), and registered in Chinese Clinical Trial Registry (registration number: ChiCtr202111090819166165).

### Participants

A total of 76 female students (aged from 18 to 30) from Beijing Sport University with shoulder MPS were enrolled in this study. The present study selected female for the reason that EIH is potentially sex-related and is more consistently observed in women^28^. The following inclusion criteria^29^ were used to choose the participants: (1) reported MPS persisting for at least 4 weeks up to 3 months. (2) had at least 1 latent trigger point on any side of the upper trapezius. The diagnosis of MPS followed the following standard: (1) palpation of a taut band, (2) identification of an exquisitely tender nodule, i.e., the myofascial trigger points in the taut band, (3) reproduction of the patient’s symptomatic pain with sustained pressure and (4) the local twitch response (LTR). The threshold value of the visual analog scale (VAS) for the MPS is set at 30mm/100mm. If multiple trigger points were detected in a participant, the trigger point with the lowest threshold of the pressure pain would be selected.

Individuals were excluded if they met the following standards: (1) confirmed or suspected spinal/shoulder injury, dislocation, fracture or inflammatory or infective diseases. (2) had a history of spinal/shoulder surgery within 12-months or other physical treatment within 1-month. (3) presentation of psychosis, depression, cognitive impairment or taking drugs for anti-depressant or anti-convulsant treatments, (4) workout with isometric, isotonic and PNF training during the experiments.

All participants were randomly allocated into 4 groups: group A (isometric exercise), group B (isotonic exercise), group C (PNF exercise) and group D (control), the randomized sequences were generated by computer. The screeners of the participants were AN and XZH.

### Procedures

This study is designed as a randomized controlled trial. Participants who were included in this study were invited to perform exercise interventions of either isometric (group A), isotonic (group B) or PNF (group C) exercises, while the participants in control group D would keep rest for 15min during the intervention session. Each exercise consists 2 scapula movements and 1 shoulder movement. The CPM and EIH responses (measured by PPT of trigger point and other remote limbs) were assessed before and after exercise session as outcome measurements. The VAS, height, weight and duration of shoulder pain were also collected before the intervention. After the outcome measure and statistical analysis, all of the participants received compensatory exercise and manual treatments. (Figure 1)

**Fig. 1.**
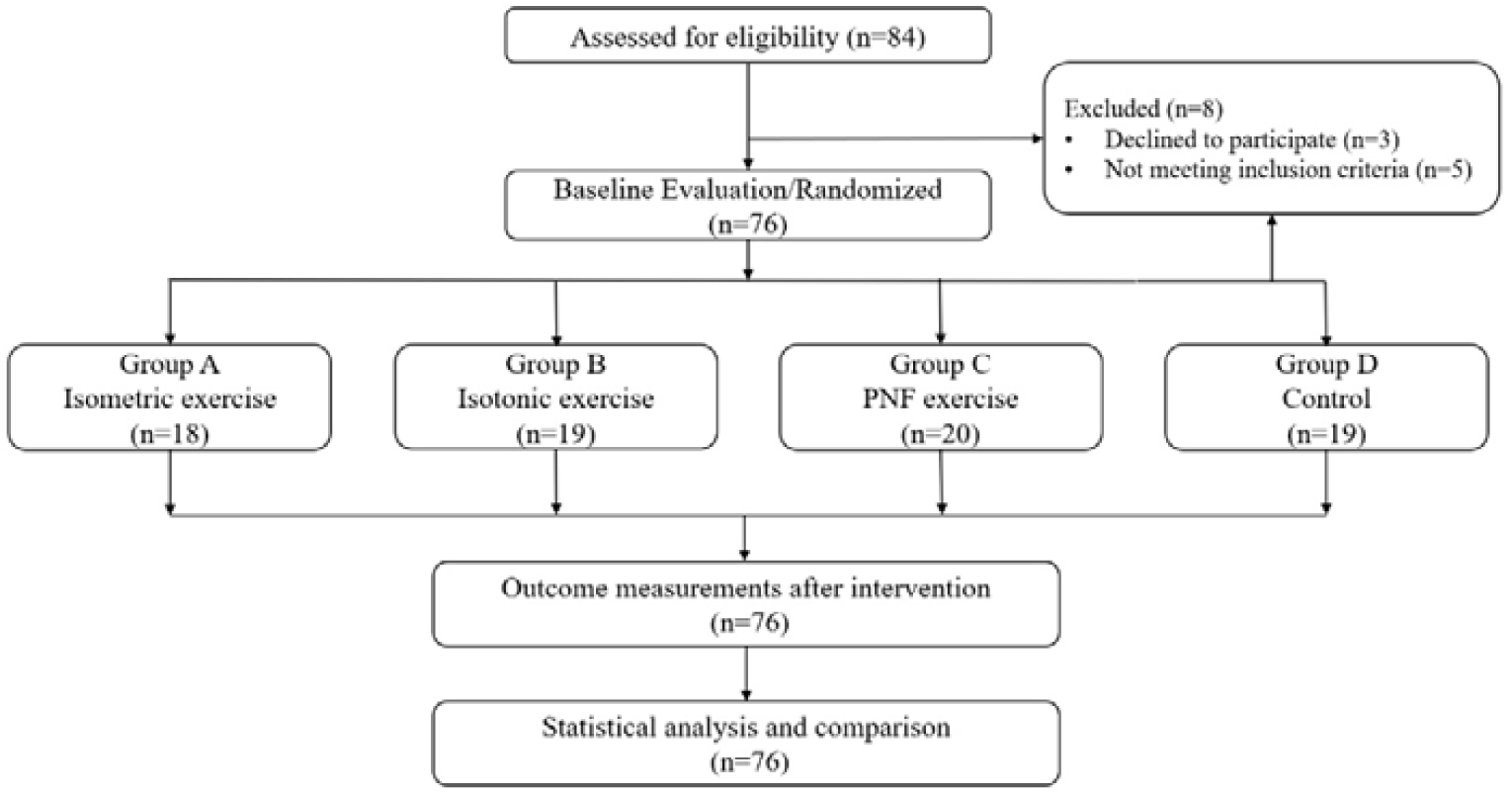
Flowchart of intervention

### Interventions

#### Isometric exercise

Participants in group A performed a modified isometric exercise program^30^ including 2 scapula movements of scapula retraction (arm row at neutral position) and scapula elevation (dumbbell shrug at neutral position), and 1 shoulder abduction, where the dumbbells with adjustable weights were used. Scapula retraction and elevation was performed at 60% maximum voluntary contraction (MVC) or pain-free load for 10s with 15 sets at neutral position. The shoulder isometric abduction was performed at 90° and the elbow was flexed at 90°, with 60% MVC for 10s of holding / 15 sets, and 30s of rest was given between sets.

The MVCs of each movement were measured by a tension dynamometer with an LCD screen providing real-time statistics, where the participants were asked to perform a set of maximum voluntary contraction at neutral position, and adjust the contraction intensities by themselves during the exercise sessions. At 1 week before the intervention, this procedure was performed three times with a 30s interval, and the mean value was identified as MVC.

#### Isotonic exercise

Participants in group B performed a modified isotonic exercise program^31,32^ including 2 scapula movements of dumbbell shrug and arm row, and 1 shoulder lateral raise, where the dumbbells with adjustable weights were used. All of the isotonic exercises were performed at moderate intensity^33^ of 60% MVC or pain-free load for 10 repetitions/5 sets, where 1 min of rest was given between sets.

#### PNF exercise

Participants in group C performed a modified PNF exercise program^34,35^ including the Intergration of Agonist Reversals (AR), Combination of Isotonic Contraction (CI), and Rhythmic Stabilization (RS) technique with the scapular pattern and upper extremity pattern. The upper extremity pattern D2 (flexion, abduction, and external rotation) was carried out using CI; the AR followed by RS was assigned to the scapular pattern D2 (anterior descending and posterior evaluation). All of the PNF exercise sessions performed 10 repetitions/5 sets at approximately 60% MVC or pain-free load, with 1 min of rest between sets.

### Outcome measures

#### PPTs of Trigger points

PPT were measured by a quantitative sensory testing protocol^36^ via handheld pressure algometer (Baseline Dolorimeter, Fabrication Enterprises, USA) with a 1 cm^2^ metal probe and applied at a rate of 0.5kg/s. PPT was measured in the trigger point located in the upper trapezius, which was labeled by sterile marker. Participants were instructed to report as soon as they perceived a pain intensity by VAS score 40 out of 100 (Pain40) during pressure application, then that threshold was recorded as PPT. This test performed 1 min before and after intervention, while the difference of PPTs during the exercise session was recorded as the local EIH responses.

#### PPTs of remote sites

The PPTs were measured at the extensor carpus radialis (test point of arms) and the peroneus longus (test point of legs) ipsilateral to the exercise limbs, and performed 1 min before and after each exercise session. The difference of PPTs during the exercise session was recorded as the remote EIH responses.

#### CPM

The CPM response was measured by quantitative sensory testing protocol^37^, with the test stimulation applied by pressure and conditioned stimulation applied by cold water immerse^38^. Participants first receive pressure stimulation at the ipsilateral extensor carpus radialis and report a PPT at Pain40 as a test stimulus. Then participants were instructed to immerse the contralateral hand into cold water at 8LJ for 1min, and report the PPT at Pain40 when the pressure applied again at the threshold after 30s of immersing. The difference between the two PPTs was recorded as the response of CPM.

#### VAS

VAS was measured using a scale printed with a line ranging from 0mm (no pain) to 100mm (worst pain), and participants were asked to locate a point on the line to rate the current pain intensity.

### Data analysis

The main outcome of this study was the PPT of trigger point, while the secondary outcomes were the PPT of remote site and CPM responses. Normality of all data was assessed by means of the 1-sample Kolmogorov-Smirnov test. Difference of baseline data (height, weight, duration of pain and VAS) between groups was verified by the 1-way ANOVA test.

The 1-way ANCOVA was used to examine whether there was a significant difference within 4 groups considering PPT, EIH and CPM at post-exercise, while the pre-exercise measurements were set as covariates. Bonferroni method was applied in the post-hoc multiple comparison. The paired t-test was used for the comparison within groups. All data were processed using SPSS

Version 21.0, and the statistical significance was set at p<0.05 for all tests.

## Results

### Baseline characteristics

Of the 76 participants with MPS, 18 of them in group A completed isometric exercise, 19 of them in group B completed isotonic exercise, 20 of group C completed PNF exercise and 19 of group D finished blank session. The average duration of shoulder MPS among participants was 7.56, 6.53,

6.85 and 7.53 weeks in group A, B, C, and D, respectively. Majority of participants (63.1%) with MPS presented the duration greater than 6 weeks, and 32.9% of them indicated over 8 weeks. Prior to the first exercise intervention, over half of the participants (57.9%) were suffering moderate to severe pain syndrome, with a VAS score higher than 40mm/100mm. All baseline participant characteristics, including age (p=0.954), height (p=0.610), weight (p=0.878), duration of pain (p=0.538) and VAS (p = 0.184), did not present significant differences between the groups (Table 1).

**Table 1.** Baseline characteristic (M±SD) *

| Measurements | A (n=18) | B(n=19) | C(n=20) | D(n=19) | P |
| --- | --- | --- | --- | --- | --- |
| Age (year) | 21.38±2.50 | 20.84±1.92 | 21.00±1.81 | 21.21±2.41 | 0.954 |
| Height (cm) | 164.83±3.75 | 166.00±4.53 | 164.55±4.21 | 166.21±4.44 | 0.610 |
| Weight (kg) | 55.03±7.27 | 57.32±9.31 | 54.75±5.37 | 59.57±8.17 | 0.878 |
| Duration of pain (week) | 7.56±2.83 | 6.53±2.95 | 6.85±2.52 | 7.53±3.01 | 0.538 |
| VAS (mm) | 45.08±12.80 | 43.76±15.89 | 43.67±13.78 | 42.63±8.06 | 0.184 |

\*: 1-way ANOVA, significant difference was set by $p<0.05$ .

### The EIH and CPM following exercises

There was a significant increase in PPT at trigger point and arm site after isotonic (p<0.000) and PNF exercises (p<0.000) while the isometric exercise and control group showed no difference compared to the baseline. The PPT at the arm sites significantly improved following PNF(p<0.000) and isotonic exercises(p<0.000), and the PPT at the leg sites also changed after isometric (p=0.025), isotonic(p=0.030) and PNF (p<0.000) exercises.

A single session of isotonic (p=0.011) and PNF (p=0.001) exercises significantly improved the CPM responses, while the isometric exercise and control group showed no difference compared to the baseline (Table 2 and Figure 2).

**Table 2.**
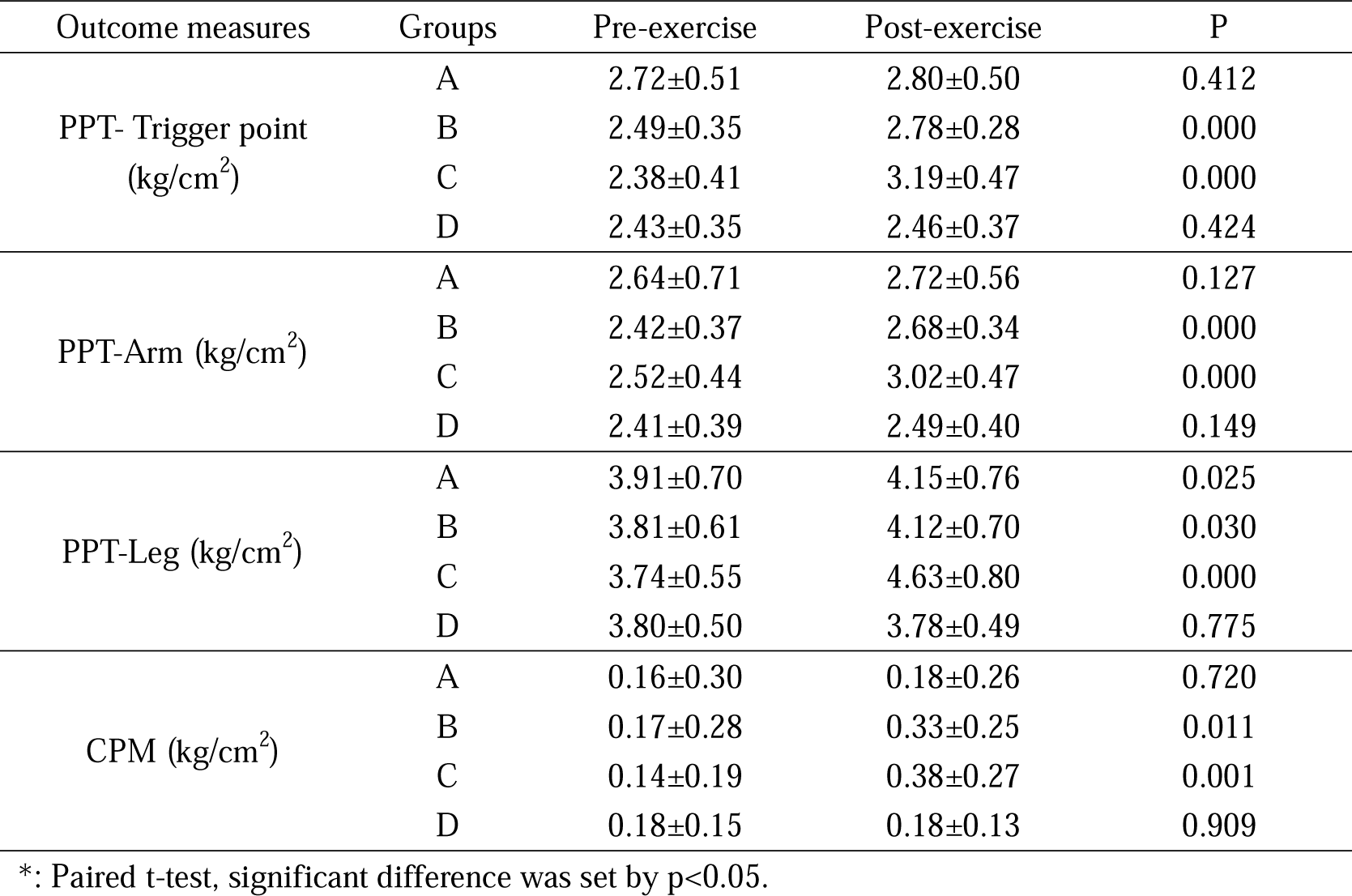
Intervention results within groups (M±SD) *

| Outcome measures | Groups | Pre-exercise | Post-exercise | P |
| --- | --- | --- | --- | --- |
| PPT- Trigger point<br>(kg/cm<sup>2</sup>) | A | 2.72±0.51 | 2.80±0.50 | 0.412 |
|  | B | 2.49±0.35 | 2.78±0.28 | 0.000 |
|  | C | 2.38±0.41 | 3.19±0.47 | 0.000 |
|  | D | 2.43±0.35 | 2.46±0.37 | 0.424 |
| PPT-Arm (kg/cm<sup>2</sup>) | A | 2.64±0.71 | 2.72±0.56 | 0.127 |
|  | B | 2.42±0.37 | 2.68±0.34 | 0.000 |
|  | C | 2.52±0.44 | 3.02±0.47 | 0.000 |
|  | D | 2.41±0.39 | 2.49±0.40 | 0.149 |
| PPT-Leg (kg/cm<sup>2</sup>) | A | 3.91±0.70 | 4.15±0.76 | 0.025 |
|  | B | 3.81±0.61 | 4.12±0.70 | 0.030 |
|  | C | 3.74±0.55 | 4.63±0.80 | 0.000 |
|  | D | 3.80±0.50 | 3.78±0.49 | 0.775 |
| CPM (kg/cm<sup>2</sup>) | A | 0.16±0.30 | 0.18±0.26 | 0.720 |
|  | B | 0.17±0.28 | 0.33±0.25 | 0.011 |
|  | C | 0.14±0.19 | 0.38±0.27 | 0.001 |
|  | D | 0.18±0.15 | 0.18±0.13 | 0.909 |

\*: Paired t-test, significant difference was set by $p<0.05$ .

**Fig. 2.**
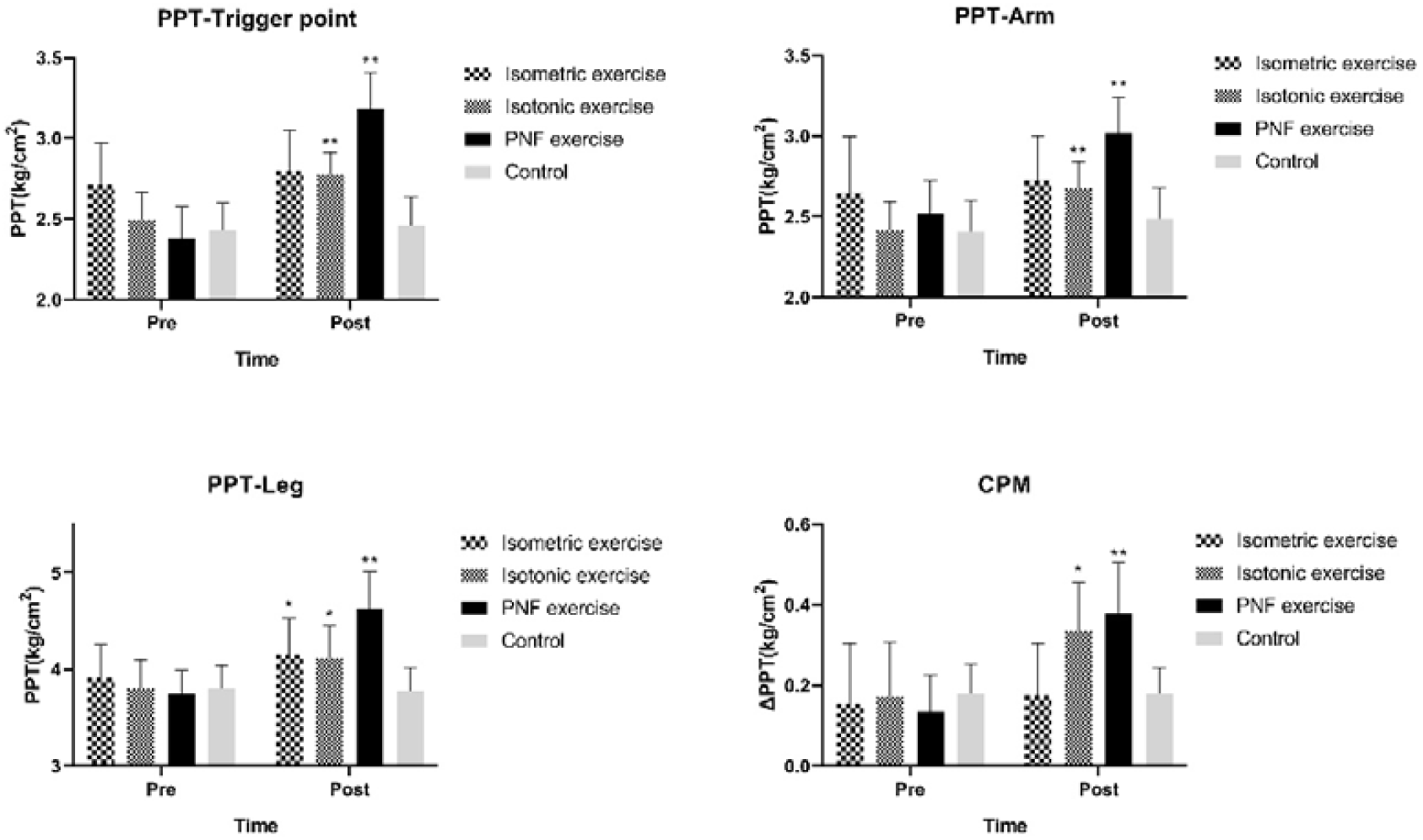
Results of PPT, EIH and CPM following exercise interventions. *=significant changes (p<0.05); **=very significant changes (p<0.01)

### The effect of exercise type on EIH and CPM

For the PPT of trigger point site, both the PNF and isotonic exercises showed significantly higher increasement compared to control group. Meanwhile, the effect of PNF exercise was greater than isotonic and isometric exercises, and the isotonic was higher than the isometric exercises. (Table 3)

**Table 3.** Between-group comparison results of the intervention*

| Outcome Measures | Groups |  | Mean Difference | Std. Error | Sig. | 95% CI |  |
| --- | --- | --- | --- | --- | --- | --- | --- |
|  |  |  |  |  |  | Lower Bound | Upper Bound |
| PPT-Trigger point (kg/cm<sup>2</sup>) | A | B | -0.144 | 0.093 | 0.764 | -0.397 | 0.109 |
|  |  | C | -0.638* | 0.094 | 0.000 | -0.894 | -0.383 |
|  |  | D | 0.130 | 0.094 | 1.000 | -0.126 | 0.385 |
|  | B | C | -0.495* | 0.090 | 0.000 | -0.734 | -0.243 |
|  |  | D | 0.274* | 0.090 | 0.021 | 0.028 | 0.519 |
|  | C | D | 0.768* | 0.089 | 0.000 | 0.526 | 1.010 |
| PPT-Arm (kg/cm<sup>2</sup>) | A | B | -0.123 | 0.086 | 0.953 | -0.351 | 0.111 |
|  |  | C | -0.389* | 0.085 | 0.000 | -0.618 | -0.160 |
|  |  | D | 0.060 | 0.086 | 1.000 | -0.174 | 0.294 |
|  | B | C | -0.266* | 0.083 | 0.012 | -0.492 | -0.040 |
|  |  | D | 0.183 | 0.084 | 0.197 | -0.045 | 0.411 |
|  | C | D | 0.449* | 0.083 | 0.000 | 0.223 | 0.675 |
| PPT-Leg (kg/cm<sup>2</sup>) | A | B | -0.053 | 0.165 | 1.000 | -0.500 | 0.395 |
|  |  | C | -0.623* | 0.163 | 0.002 | -1.067 | -0.180 |
|  |  | D | 0.285 | 0.165 | 0.529 | -0.162 | 0.733 |
|  | B | C | -0.570* | 0.160 | 0.004 | -1.006 | -0.135 |
|  |  | D | 0.338 | 0.162 | 0.246 | -0.103 | 0.779 |
|  | C | D | 0.908* | 0.160 | 0.000 | 0.473 | 1.344 |
| CPM (kg/cm<sup>2</sup>) | A | B | -0.152 | 0.069 | 0.186 | -0.339 | 0.034 |
|  |  | C | -0.212* | 0.068 | 0.016 | -0.397 | -0.027 |
|  |  | D | 0.007 | 0.069 | 1.000 | -0.181 | 0.194 |
|  | B | C | -0.061 | 0.067 | 1.000 | -0.243 | 0.122 |
|  |  | D | 0.159 | 0.068 | 0.135 | -0.026 | 0.344 |
|  | C | D | 0.219* | 0.067 | 0.011 | 0.036 | 0.402 |

\*: 1-way ANCOVA, adjusted by Bonferroni. significant difference was set by p<0.05.

And for the PPT of remote sites, only the PNF exercises increased significantly higher thresholds compared to the control group and the isometric exercises at both arm and leg sites. However, the isotonic exercise also had a greater improvement than the isometric exercise, but not with the control group. And for the CPM, only PNF exercise attained a significantly greater responses compared to both control group and isometric exercise (Table 3).

## Discussion

This pilot study investigated the local and remote responses of EIH and CPM after PNF, isotonic, and isometric resistance exercises for patients with MPS. Our findings mostly met what we previously hypothesized, where was an increase in PPT at trigger point, arm and leg sites with the CPM responses demonstrated in participants performed PNF and isotonic exercise, while the isometric exercise only increased PPT at leg sites. Compared with control group, both isotonic and PNF group showed significant greater EIH responses at the trigger points. However, only the PNF exercise significantly improved PPT at remote sites and CPM responds compared to the control group.

MPS, generally regarded as a typical chronic musculoskeletal pain^39^, is mainly characterized by the presence of trigger points^40^, which is a hypersensitive area that can be palpated in a muscle taut band. The trigger point is possibly induced by a continuous nociceptive stimulus from the local energy crisis of overused fibers^41^, followed by pain sensitization^42^ which means the impairment of descending pain modulation. Thus, chronic MPS could attenuate both the CPM and the EIH effect, Vaegter et al^43^ reported an altered effect of EIH in patients with chronic low back pain, where the acute 6-min walk failed to induce EIH in patients with greater pain sensitivity. Chretien et al^5^ also found that the deficits of EIH were related to the reduced CPM among adolescent girls with chronic pain.

In this study, the CPM responses of MPS patients were significantly increased following both PNF and isotonic exercise in this study. It’s believed that exercise with optimal intensity and type could affect central pain modulation, mediate neurotransmitters and cytokines. Activation of the endocannabinoids, endogenous opioids, and 5-HT system after exercises with various resistances may enhance the descending inhibition and reduce pain. Crombie et al^44^ investigated that the serum endocannabinoids increased alongside attenuation of pain sensitivity after resistance exercises in healthy individuals, while Bobinski et al^13^ also found an increase of 5-HT in RVM through low-intensity exercise. Meanwhile, the activation of pain-related cortex regions following exercises may also mediate pain processing of the thalamus and PAG. Cummiford et al^45^ found that the pain perception and facilitation of the thalamus was confined by M1 stimulation, while Ellingson et al^46^ reported that the DLPFC function improved and was correlated with pain reduction in patients with fibromyalgia after cycling exercise. Interestingly, Lial et al^47^ found that PNF exercise significantly improved DLPFC activation, which may have potential effects on the central pain modulation, and still need further investigation.

In this study, EIH was demonstrated at the trigger point, arm and leg sites for MPS patients following PNF and isotonic exercises, which is consistent with prior research under various painful conditions. First, the global analgesic effects induced by PNF and resistance exercises were also verified by Burrows et al^23^, that the isotonic shoulder exercises of non-painful limbs in patients with knee osteoarthritis successfully reduced pain, while Kuppens et al^48^ confirmed EIH responses at leg sites after moderate intensity shoulder extension exercises. Koltyn er al^49^ also demonstrated the EIH responses elicited by contralateral isometric contraction in healthy individuals. Second, the isotonic and PNF exercises elicited greater EIH than isometric exercises, which is also found by Chung et al^50^ that the isotonic exercises have better EIH responses in patients with chronic neck pain than isometric exercises. Lastly, only isometric exercise failed to attenuate the pain perception of trigger point, which is also consistent with a previous systematic review by Bonello et al^51^ indicating that there is no consistent evidence for EIH following isometric exercises in chronic pain patients, and Staud et al^52^ also found that isometric exercise increased pain intensity in patients with fibromyalgia.

Neuromuscular exercises such as PNF have a significant therapeutic effect on many musculoskeletal pain conditions. A Meta-analysis by Gao et al^18^ showed that PNF have more beneficial effects in pain relief and waist function improvement in patients with chronic LBP compared with other exercise intervention. Regarding chronic neck pain, Lytras et al^53^evaluated neuromuscular inhibition therapy combined with exercise intervention, the results showed that the combined program successfully reduced pain rating and improved neck function, while PNF also has a significant effect on knee osteoarthritis^54^ and patellofemoral pain syndrome^55^.

In this study, PNF exercise had a greater analgesic effect on MPS after intervention compared to control group and other exercises, which may result from the enhanced proprioception and C fibers inputs from the additional eccentric^34^ and dynamic muscle contractions. Although the activation of noxious C fibers following the maximum eccentric exercise^19^ may trigger the mechanical allodynia^56^ and delayed onset muscle soreness, the sufficient noxious or non-noxious C fibers input during the sub-maximum eccentric in PNF or even the isotonic exercise may still activate the decsending inhibition via thalamus VM nucleus. Stackhouse et al^57^ compared the analgesic effect between the noxious electrical stimulation and the eccentric plantar flexor exercise with moderate intensity, found that eccentric exercise induced both mechanical and thermal pain perception effectively.

It’s concerned that only the isotonic exercise showed a greater change of PPT compared to control group at the trigger point besides from the PNF exercise, which suggested that the onset hypoalgesia affect from the CPM test might contributed these changes during the test. The CPM test applied in this study which executed cold stimulus can also activate the C-fiber afferent and the descending inhibition, which may have the overlap effect with the EIH responds. Thus, the relationship between the CPM test and the exercise still needs further investigation.

This study has several limitations. First, the indicators of the pain tests were limited, for instance, the PPT combined with heat pain thresholds might better reflect the real neurophysiological characteristics of musculoskeletal pain. Second, individuals with MPS were diversified by pain duration and intensity, which may affect the consistency of the results. Third, all of the participants were female, so the possible gender difference of descending pain processing should be considered in future studies. Lastly, the EIH efficiency of moderate intensity resistance exercise in this work was insufficient investigated and thus still needs to be evaluated comprehensively by increasing the variety of exercise types and duration length of the interventions.

## Conclusions

In summary, PNF, isotonic and isometric exercises could exert significant local and global EIH effects for MPS patients, which could be affected by the proprioception stimulus under the exercise types. The significant increases in CPM response after PNF and isotonic exercises indicated that the EIH mechanisms of these moderate intensity exercises may involve the enhancement of the central descending inhibitory function. The findings of this study can provide theoretical references to further studies focusing on central mechanisms of EIH, which could optimize the effect of exercise interventions for chronic pain in future clinical practices.

## Data Availability

All data produced in the present study are available upon reasonable request to the authors

## Acknowledgments

We would like to thank He-Jing Xu, Yang-Zhi Wu, Xi-Min Wu, Zi-Hang Gao, Wei Liu, Mao-Li Li, Li-Hui Qin, Zhen-Yan Chen, Cheng-Huan Hao, Bai-Hong Meng and all the researchers who provided help and advice in our experiments.

## References

1. Sitges C, Velasco-Roldan O, Crespi J, et al. Acute Effects of a Brief Physical Exercise Intervention on Somatosensory Perception, Lumbar Strength, and Flexibility in Patients with Nonspecific Chronic Low-Back Pain. J Pain Res. 2021;14:487–500.

2. Wewege MA, Jones MD. Exercise-Induced Hypoalgesia in Healthy Individuals and People With Chronic Musculoskeletal Pain: A Systematic Review and Meta-Analysis. J Pain. 2021;22(1):21–31.

3. Holmes SA, Kim A, Borsook D. The brain and behavioral correlates of motor-related analgesia (MRA). Neurobiol Dis. 2021;148:105158.

4. Shah JP, Thaker N, Heimur J, Aredo JV, Sikdar S, Gerber L. Myofascial Trigger Points Then and Now: A Historical and Scientific Perspective. PM R. 2015;7(7):746–761.

5. Chretien R, Lavoie S, Chalaye P, et al. Reduced endogenous pain inhibition in adolescent girls with chronic pain. Scand J Pain. 2018;18(4):711–717.

6. Kuithan P, Heneghan NR, Rushton A, Sanderson A, Falla D. Lack of Exercise-Induced Hypoalgesia to Repetitive Back Movement in People with Chronic Low Back Pain. Pain Pract. 2019;19(7):740–750.

7. You HJ, Lei J, Niu N, et al. Specific thalamic nuclei function as novel ‘nociceptive discriminators’ in the endogenous control of nociception in rats. Neuroscience. 2013;232:53–63.

8. You HJ, Lei J, Pertovaara A. Thalamus: The ‘promoter’ of endogenous modulation of pain and potential therapeutic target in pathological pain. Neurosci Biobehav Rev. 2022;139:104745.

9. Lei J, Sun T, Lumb BM, You HJ. Roles of the periaqueductal gray in descending facilitatory and inhibitory controls of intramuscular hypertonic saline induced muscle nociception. Exp Neurol. 2014;257:88–94.

10. Sagalajev B, Viisanen H, Wei H, Pertovaara A. Descending antinociception induced by secondary somatosensory cortex stimulation in experimental neuropathy: role of the medullospinal serotonergic pathway. J Neurophysiol. 2017;117(3):1200–1214.

11. Da Silva LF, Walder RY, Davidson BL, Wilson SP, Sluka KA. Changes in expression of NMDA-NR1 receptor subunits in the rostral ventromedial medulla modulate pain behaviors. Pain. 2010;151(1):155–161.

12. Kim YJ, Byun JH, Choi IS. Effect of Exercise on micro-Opioid Receptor Expression in the Rostral Ventromedial Medulla in Neuropathic Pain Rat Model. Ann Rehabil Med. 2015;39(3):331–339.

13. Bobinski F, Ferreira TAA, Cordova MM, et al. Role of brainstem serotonin in analgesia produced by low-intensity exercise on neuropathic pain after sciatic nerve injury in mice. Pain. 2015;156(12):2595–2606.

14. Taylor BK, Westlund KN. The noradrenergic locus coeruleus as a chronic pain generator. J Neurosci Res. 2017;95(6):1336–1346.

15. Borovskis J, Cavaleri R, Blackstock F, Summers SJ. Transcranial Direct Current Stimulation Accelerates The Onset of Exercise-Induced Hypoalgesia: A Randomized Controlled Study. J Pain. 2021;22(3):263–274.

16. Zhou YS, Meng FC, Cui Y, et al. Regular Aerobic Exercise Attenuates Pain and Anxiety in Mice by Restoring Serotonin-Modulated Synaptic Plasticity in the Anterior Cingulate Cortex. Med Sci Sports Exerc. 2022;54(4):566–581.

17. Zhang X, Zong B, Zhao W, Li L. Effects of Mind-Body Exercise on Brain Structure and Function: A Systematic Review on MRI Studies. Brain Sci. 2021;11(2).

18. Gao P, Tang F, Liu W, Mo Y. The effects of proprioceptive neuromuscular facilitation in treating chronic low back pain: A systematic review and meta-analysis. J Back Musculoskelet Rehabil. 2021.

19. Adreani CM, Hill JM, Kaufman MP. Responses of group III and IV muscle afferents to dynamic exercise. J Appl Physiol (1985). 1997;82(6):1811–1817.

20. Areeudomwong P, Buttagat V. Proprioceptive neuromuscular facilitation training improves pain-related and balance outcomes in working-age patients with chronic low back pain: a randomized controlled trial. Braz J Phys Ther. 2019;23(5):428–436.

21. Sipko T, Glibowski E, Kuczynski M. Acute effects of proprioceptive neuromuscular facilitation exercises on the postural strategy in patients with chronic low back pain. Complement Ther Clin Pract. 2021;44:101439.

22. Lannersten L, Kosek E. Dysfunction of endogenous pain inhibition during exercise with painful muscles in patients with shoulder myalgia and fibromyalgia. Pain. 2010;151(1):77–86.

23. Burrows NJ, Booth J, Sturnieks DL, Barry BK. Acute resistance exercise and pressure pain sensitivity in knee osteoarthritis: a randomised crossover trial. Osteoarthritis Cartilage. 2014;22(3):407–414.

24. Ramaswamy S, Wodehouse T. Conditioned pain modulation-A comprehensive review. Neurophysiol Clin. 2021;51(3):197–208.

25. Vidor LP, Torres IL, Medeiros LF, et al. Association of anxiety with intracortical inhibition and descending pain modulation in chronic myofascial pain syndrome. BMC Neurosci. 2014;15:42.

26. Fingleton C, Smart KM, Doody CM. Exercise-induced Hypoalgesia in People With Knee Osteoarthritis With Normal and Abnormal Conditioned Pain Modulation. Clin J Pain. 2017;33(5):395–404.

27. Vaegter HB, Handberg G, Graven-Nielsen T. Similarities between exercise-induced hypoalgesia and conditioned pain modulation in humans. Pain. 2014;155(1):158–167.

28. Naugle KM, Naugle KE, Fillingim RB, Riley JL, 3rd. Isometric exercise as a test of pain modulation: effects of experimental pain test, psychological variables, and sex. Pain Med. 2014;15(4):692–701.

29. Fernandez-de-Las-Penas C, Dommerholt J. International Consensus on Diagnostic Criteria and Clinical Considerations of Myofascial Trigger Points: A Delphi Study. Pain Med. 2018;19(1):142–150.

30. Price J, Rushton A, Tyros I, Tyros V, Heneghan NR. Effectiveness and optimal dosage of exercise training for chronic non-specific neck pain: A systematic review with a narrative synthesis. PLoS One. 2020;15(6):e0234511.

31. Dube MO, Desmeules F, Lewis J, Roy JS. Rotator cuff-related shoulder pain: does the type of exercise influence the outcomes? Protocol of a randomised controlled trial. BMJ Open. 2020;10(11):e039976.

32. Andersen LL, Kjaer M, Sogaard K, Hansen L, Kryger AI, Sjogaard G. Effect of two contrasting types of physical exercise on chronic neck muscle pain. Arthritis Rheum. 2008;59(1):84–91.

33. da Cunha Ribeiro RP, Franco TC, Pinto AJ, et al. Prescribed Versus Preferred Intensity Resistance Exercise in Fibromyalgia Pain. Front Physiol. 2018;9:1097.

34. Oh DG, Yoo KT. The effects of therapeutic exercise using PNF on the size of calcium deposits, pain self-awareness, and shoulder joint function in a calcific tendinitis patient: a case study. J Phys Ther Sci. 2017;29(1):163–167.

35. Lee JH, Park SJ, Na SS. The effect of proprioceptive neuromuscular facilitation therapy on pain and function. J Phys Ther Sci. 2013;25(6):713–716.

36. Wytrazek M, Huber J, Lipiec J, Kulczyk A. Evaluation of palpation, pressure algometry, and electromyography for monitoring trigger points in young participants. J Manipulative Physiol Ther. 2015;38(3):232–243.

37. Mertens MG, Hermans L, Crombez G, et al. Comparison of five conditioned pain modulation paradigms and influencing personal factors in healthy adults. Eur J Pain. 2021;25(1):243–256.

38. Coulombe-Leveque A, Tousignant-Laflamme Y, Leonard G, Marchand S. The effect of conditioning stimulus intensity on conditioned pain modulation (CPM) hypoalgesia. Can J Pain. 2021;5(1):22–29.

39. Saxena A, Chansoria M, Tomar G, Kumar A. Myofascial pain syndrome: an overview. J Pain Palliat Care Pharmacother. 2015;29(1):16–21.

40. Gerwin RD. Myofascial Trigger Point Pain Syndromes. Semin Neurol. 2016;36(5):469–473.

41. Simons DG. New views of myofascial trigger points: etiology and diagnosis. Arch Phys Med Rehabil. 2008;89(1):157–159.

42. Woolf CJ. Central sensitization: implications for the diagnosis and treatment of pain. Pain. 2011;152(3 Suppl):S2–15.

43. Vaegter HB, Petersen KK, Sjodsholm LV, Schou P, Andersen MB, Graven-Nielsen T. Impaired exercise-induced hypoalgesia in individuals reporting an increase in low back pain during acute exercise. Eur J Pain. 2021;25(5):1053–1063.

44. Crombie KM, Brellenthin AG, Hillard CJ, Koltyn KF. Endocannabinoid and Opioid System Interactions in Exercise-Induced Hypoalgesia. Pain Med. 2018;19(1):118–123.

45. Cummiford CM, Nascimento TD, Foerster BR, et al. Changes in resting state functional connectivity after repetitive transcranial direct current stimulation applied to motor cortex in fibromyalgia patients. Arthritis Res Ther. 2016;18:40.

46. Ellingson LD, Stegner AJ, Schwabacher IJ, Koltyn KF, Cook DB. Exercise Strengthens Central Nervous System Modulation of Pain in Fibromyalgia. Brain Sci. 2016;6(1).

47. Lial L, Moreira R, Correia L, et al. Proprioceptive neuromuscular facilitation increases alpha absolute power in the dorsolateral prefrontal cortex and superior parietal cortex. Somatosens Mot Res. 2017;34(3):204–212.

48. Kuppens K, Struyf F, Nijs J, et al. Exercise- and Stress-Induced Hypoalgesia in Musicians with and without Shoulder Pain: A Randomized Controlled Crossover Study. Pain Physician. 2016;19(2):59–68.

49. Koltyn KF, Umeda M. Contralateral attenuation of pain after short-duration submaximal isometric exercise. J Pain. 2007;8(11):887–892.

50. Chung S, Jeong YG. Effects of the craniocervical flexion and isometric neck exercise compared in patients with chronic neck pain: A randomized controlled trial. Physiother Theory Pract. 2018;34(12):916–925.

51. Bonello C, Girdwood M, De Souza K, et al. Does isometric exercise result in exercise induced hypoalgesia in people with local musculoskeletal pain? A systematic review. Phys Ther Sport. 2020;49:51–61.

52. Staud R, Robinson ME, Price DD. Isometric exercise has opposite effects on central pain mechanisms in fibromyalgia patients compared to normal controls. Pain. 2005;118(1-2):176–184.

53. Lytras DE, Sykaras EI, Christoulas KI, Myrogiannis IS, Kellis E. Effects of Exercise and an Integrated Neuromuscular Inhibition Technique Program in the Management of Chronic Mechanical Neck Pain: A Randomized Controlled Trial. J Manipulative Physiol Ther. 2020;43(2):100–113.

54. Song Q, Shen P, Mao M, Sun W, Zhang C, Li L. Proprioceptive neuromuscular facilitation improves pain and descending mechanics among elderly with knee osteoarthritis. Scand J Med Sci Sports. 2020;30(9):1655–1663.

55. Motealleh A, Mohamadi M, Moghadam MB, Nejati N, Arjang N, Ebrahimi N. Effects of Core Neuromuscular Training on Pain, Balance, and Functional Performance in Women With Patellofemoral Pain Syndrome: A Clinical Trial. J Chiropr Med. 2019;18(1):9–18.

56. Nagi SS, Mahns DA. C-tactile fibers contribute to cutaneous allodynia after eccentric exercise. J Pain. 2013;14(5):538–548.

57. Stackhouse SK, Taylor CM, Eckenrode BJ, Stuck E, Davey H. Effects of Noxious Electrical Stimulation and Eccentric Exercise on Pain Sensitivity in Asymptomatic Individuals. PM R. 2016;8(5):415–424.

